# Local and global mortality experience: A novel hierarchical model for regional mortality risk

**DOI:** 10.1101/2024.10.17.24315673

**Authors:** Asmik Nalmpatian, Christian Heumann, Levent Alkaya, William Jackson

**Affiliations:** Department of Statistics, LMU Munich, Munich, Bavaria, Germany

## Abstract

Accurate mortality risk assessment is critical for decision-making in life insurance, healthcare, and public policy. Regional variability in mortality, driven by diverse local factors and inconsistent data availability, presents significant modeling challenges. This study introduces a novel hierarchical mortality risk model that integrates global and local data, enhancing regional mortality estimation across diverse regions. The proposed approach employs a two-stage process: first, a global Light Gradient Boosting Machine model is trained on globally shared features; second, region-specific models are developed to incorporate local characteristics. This framework outperforms both purely local models and standard imputation techniques, particularly in data-scarce regions, by leveraging global patterns to improve generalization. The model is computationally efficient, scalable, and robust in handling missing values, making it adaptable for other domains requiring integration of multi-regional data. This method enhances predictive accuracy across various regions and provides a more reliable approach for mortality risk estimation in data-scarce environments.

## Introduction

Mortality risk assessment plays a crucial role in various sectors, including life insurance, healthcare, and public policy. Reliable estimates of mortality rates are essential for strategic planning, policy formulation, and ensuring the financial stability of life insurance systems. However, accurately estimating mortality risk presents an essential challenge due to the diverse and dynamic nature of regional data availability and factors that affect mortality rates.

Hierarchical models have been utilized in mortality studies to account for variations at different levels, including regional, individual and national. Originally developed in fields like education, sociology, and demography, these models have gained significant traction in public health and epidemiology. By generalizing the classical pooling of group estimates, hierarchical or multilevel models offer a flexible framework for analyzing mortality data [50]. This flexibility allows researchers to better understand and interpret the complex factors influencing mortality rates across different populations.

Existing models in hierarchical mortality modeling include Bayesian approaches, generalized linear models, and machine learning (ML) techniques. Bayesian hierarchical models estimate mortality rates by incorporating prior distributions to handle uncertainty [48]. Generalized linear models, including multilevel Poisson regression, have been applied to mortality data to account for overdispersion and hierarchical structure [49]. Although the existing literature predominantly employs random effects for both methodologies, our approach diverges by sequentially processing the residuals. Recent studies have also explored ML methods such as random forests and gradient boosting for COVID-19 mortality modeling [58].

Studies have highlighted the importance of balancing global patterns with local specifics in mortality modeling to ensure both generalizability and relevance [56, 57]. However, the availability of mortality data varies widely across regions, posing challenges for model accuracy and reliability [54]. Poisson regression is commonly used for modeling count data, including mortality rates [47], whereas Light Gradient Boosting Machine (LightGBM) has been recognized for its efficiency and accuracy in handling large datasets, making it suitable for hierarchical mortality modeling [52].

Existing mortality models often struggle to balance global trends and local variations, leading to models that either overgeneralize or fail to capture region-specific nuances. Furthermore, inconsistent and sparse data availability across regions intensifies these challenges, reducing the reliability of predictions, especially in data-scarce environments [54]. Current approaches often suffer from overdispersion [46] or are computationally inefficient when handling large datasets [53] or missing data [53]. These limitations underscore the need for a more flexible and scalable solution.

To address these challenges, this study introduces a novel hierarchical mortality modeling approach that integrates both global and local data. By using a two-stage process, our model first captures global patterns through a LightGBM model with a Poisson regression objective and then refines these predictions with region-specific models that account for local characteristics. While the first step includes shared variables that apply to all countries, such as age and gender, the country-specific models capture unique regional characteristics by incorporating additional region-specific factors, such as lifestyle habits and environmental conditions. This method markedly improves predictive performance, particularly in data-sparse regions, by leveraging global insights while remaining adaptable to the unique conditions of each region. Additionally, the model is computationally efficient, scalable, and capable of handling missing values, making it superior to traditional pooling methods. Beyond mortality risk estimation, this hierarchical modeling framework is applicable to other domains requiring multi-regional data integration, such as public health planning, epidemiological forecasting, and financial risk assessments. Its ability to generalize well across different regions makes it particularly valuable in scenarios where data sparsity or inconsistency is a common obstacle.

The structure of this paper is as follows: Section 2 provides a brief overview of our database and Section 3 presents our proposed methodology in detail. Section 4 examines the effectiveness of our methodology by presenting and discussing the results. Finally, Section 5 concludes by summarizing the main findings and suggesting research and industry perspectives.

## Database

Data for the study was collected in a pseudonymised form from eight different operating units of a global primary insurance company, each representing a distinct country. Data privacy regulations prohibit the disclosure of these countries’ names, keeping the focus on the technical aspects of the model evaluation and comparison, rather than on potential privacy breaches. The chosen organizations were based on two key factors: having relevant data available of high quality and representing diverse geographic regions.

The dataset includes policy data that remained active during this period, even if initially issued before the earliest year studied. In total, the dataset encompasses nearly 10 million life-years of exposure and close to 10,000 recorded insurance claims (=deaths).

The data underwent analysis in an aggregated form, grouped into *N* = 16.689.304 unique combinations of feature values. Specifically, the feature set *X*_*i,j*_, where group *i* ranges from 1 to *N* and *j* ranges from 1 to 8 -representing the eight countries, consists of a total of 26 features. Among these features, 9 are global, and up to 17 are local features, encompassing information about policyholders, insurance policies, and claims. Given these potential risk factors, our target is to model the number of deaths *D*_*i,j*_ in relation to the life years of risk exposure *E*_*i,j*_. To facilitate model training and evaluation, an artificial variable was constructed before aggregating to create an 80-20 train-test split, ensuring that all unique combinations are adequately represented in both the training and test sets.

Table 1 provides an overview of *D*_*i,j*_, *E*_*i,j*_, and the total number of years included *T*_*i,j*_ for group *i* in country *j*, thereby facilitating a comprehensive understanding of the dataset’s key characteristics and distribution.

**Table 1.** Overview of death counts *D*_*j*_, exposure in life years *E*_*j*_, unique feature combination *N*_*j*_, and observed years *T*_*j*_ for each country *j*.

| Country $j$ | $D_j$ | $E_j$ | $N_j$ | $T_j$ |
| --- | --- | --- | --- | --- |
| 1 | 1699 | 1295299 | 1880792 | 2013–2020 |
| 2 | 1291 | 1686299 | 2190943 | 2010–2020 |
| 3 | 494 | 815795 | 1868691 | 2010–2020 |
| 4 | 1225 | 1347150 | 1572539 | 2017–2020 |
| 5 | 1816 | 1825901 | 4825792 | 2016–2020 |
| 6 | 2132 | 1548157 | 3852306 | 2016–2020 |
| 7 | 458 | 498560 | 207951 | 2017–2020 |
| 8 | 297 | 99473 | 290290 | 2015–2020 |
| **Total** | **9412** | **9116634** | **16689304** | **2010–2020** |

## Methodology

The foundation of our approach is rooted in the Cox Proportional Hazards model (Cox PH), a class of survival models in statistics that aligns with our objective of estimating mortality rates [2]. To simplify the complexity of Cox PH model calculations, we leveraged the connection between Cox PH and a Poisson Generalized Linear Model (GLM). Assuming piecewise constant hazard rates over time, the likelihood of the Cox PH model coincides with the likelihood of the Poisson GLM when we employ *log*(*E*_*i,j*_) as an offset parameter, as detailed by [29] who noted, “we do not assume [the Poisson model] is true, but simply use it as a device for deriving the likelihood”. Independent of [29], [45] published a similar insight, emphasizing that the piece-wise proportional hazards model is equivalent to a specific Poisson regression model.

Our primary goal is to accurately evaluate mortality rates. We aim to estimate the conditional expectation of death counts, denoted as *D*_*i,j*_, given the available information summarized in the feature set *X*_*i,j*_ and the exposure in life years at risk *E*_*i,j*_. Assuming that 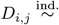 Poisson(*μ*_*i,j*_ · *E*_*i,j*_), the expectation according to the Poisson distributional assumption is:

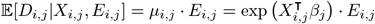

The Poisson log-likelihood is defined:

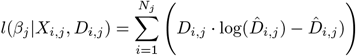

where *D*_*i,j*_ denotes the observed death counts, 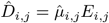 denotes the predicted death counts, and *β*_*j*_ is the parameter vector.

This formulation assumes that deaths follow a Poisson distribution. An advantage of simplifying the Cox PH model into a Poisson GLM is its adaptability to the ML realm, requiring optimization using Poisson log-likelihood and the ability to define an offset or observation weights. ML models, which generally do not assume specific (i.e. additive) relationships between features and targets, can leverage this flexibility:

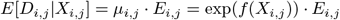

This transition from GLMs to ML models offers additional benefits, including integrated variable selection mechanisms and the ability to capture interactions without explicit specification.

To implement this approach, we employ the LightGBM algorithm [52], a popular ML technique based on boosting. LightGBM iteratively builds an ensemble of decision trees to model the relationship between features and the target variable, optimizing the model to minimize the negative log-likelihood of the Poisson distribution [25]. Trees are fit to residuals derived from the loss function, and the model is updated iteratively to minimize this loss. The prediction is formulated as a linear combination of the base learners:

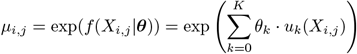

where *θ*_*k*_ is the weight of the *k*-th tree, and 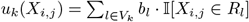 represents the tree associated with *V*_*k*_ as set of leaves of the *k*-th tree, *b*_*l*_ as the predicted value in the *l*-th leaf, and *R*_*l*_ as the region defined by disjoint partitions of the training set associated with the *l*-th leaf [28]. LightGBM uses a leaf-wise growth strategy, splitting the leaf with the highest loss reduction first, and adopts a histogram-based algorithm to improve the efficiency and speed of building decision trees. This approach results in efficient and accurate models, particularly for datasets with complex or imbalanced relationships. Mechanisms we employ to control overfitting and ensure robust performance are detailed in.

### Two-step model

To distinguish between local and global features and ensure high accuracy in each country, we propose a Two-step model approach. This approach involves two distinct modeling steps:

#### Step 1: Global model

The first model identifies global patterns and uses a training set that includes data from all countries, focusing solely on “global” factors. These global factors are those where data across countries is comparable, such as age. In contrast, factors like postal code, which lack comparability between regions, are excluded.

#### Step 2: Specialized Local model

In the second step, we calculate one Local model per country, totaling eight Local models. Each Local model takes the output of the Global model and adjusts it to the specific circumstances of the respective country. Specialized Local models use all global factors plus the country-specific local factors. The distinction of the feature set into global and local features is based on the availability of data across countries as well as domain-specific expert knowledge.

This approach combines the estimates from both the global and specialized Local models as illustrated in Fig 1.

**Fig 1.**
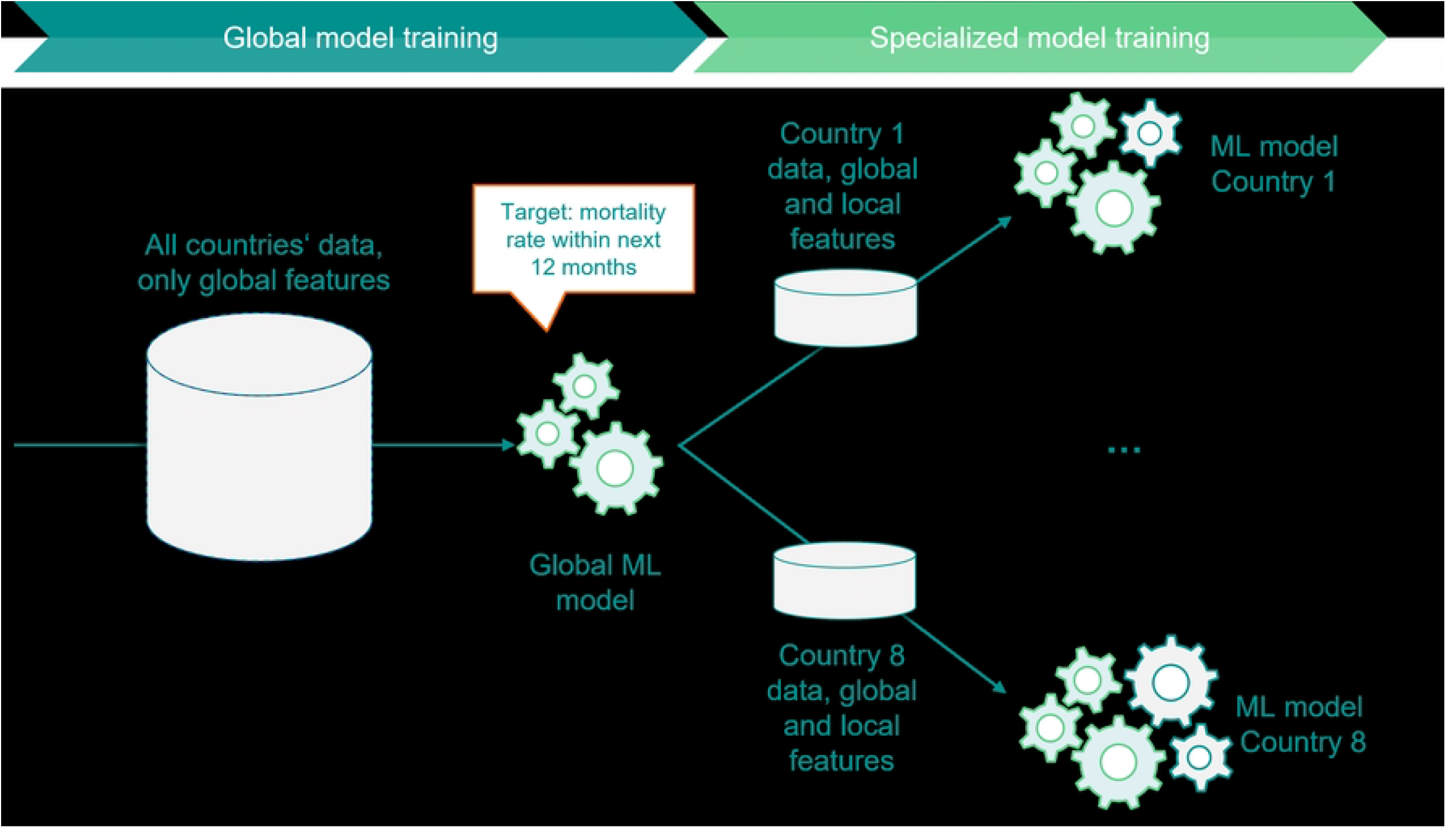
Qualitative illustration of the proposed methodology, with gearwheels representing the features.

Mathematically, we can express the process of estimating death counts for a policy with given factors as follows:

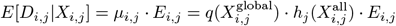

where *D*_*i,j*_ represents the expected number of deaths given a set of features *X*_*i,j*_ for group *i* and country *j*; *q*(·) represents the Global model’s prediction function; *h*_*j*_(·) represents the Local model’s prediction function for country *j*; 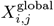 represents a set of factor values for group *i* and country *j*, containing only global factors; 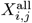 represents a set of factor values for country *j*, containing both global and local factors.

In technical terms, the predicted mortality rates from the first Global model are used to initialise the second specialized Local model. Accordingly, the model continues to work on the resulting residuals and iteratively optimises the second model - but now with the broader, localised data set. The final predicted number of deaths results from the multiplication of the predictions from the Global model (first step), the predictions from the specialised Local model (second step) and the exposure. The following derivation shows that the multiplication is justified by the nature of the boosting algorithm and the exponentiation by the log link of the Poisson distribution:

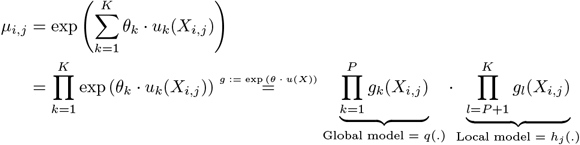

Splitting the modeling into two steps offers the advantage of cleanly separating effects into local and global categories. It also optimizes model performance for each market by tailoring the model to local patterns while allowing knowledge sharing across countries via the Global model. Additionally, when onboarding a new country, we can choose to retain the existing Global model and calculate a new Local model for this new country.

We employ Microsoft’s ML library “LightGBM” for implementing these models, which have demonstrated high accuracy in various scenarios. As the software does not allow the inclusion of an offset, we utilize observed mortality rates as the target variable, thus the death counts are scaled by exposure *D*_*i,j*_*/E*_*i,j*_ and exposure *E*_*i,j*_ is used as weights, a method demonstrated to be mathematically equivalent in the Poisson case by [33]. These residuals *R*_*i,j*_ represent the deviation of the observed deaths from the expected deaths 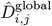 predicted by the first step, and are calculated as follows: 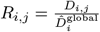. In the second step, these residuals serve as the target variable for further modeling. The new weights for this step are the expected deaths from the first step, 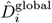. It is important to note that in the second step, we use the complete feature set of a single country, whereas in the first step, we utilize pooled data with global features only. Details on prediction calibration are provided in.

## Benchmarking results

Our objective is to benchmark the proposed methodology against three other approaches using specific evaluation metrics. This aims to determine the predictive performance and computational efficiency of the proposed model compared to the alternatives. All these methods are based on the model specification proposed in the previous section, where death counts are estimated in relation to exposure using the ML model LightGBM, optimizing the Poisson log-likelihood assumption. The differences among these methods are outlined below and illustrated in Fig 2:

**Fig 2.**
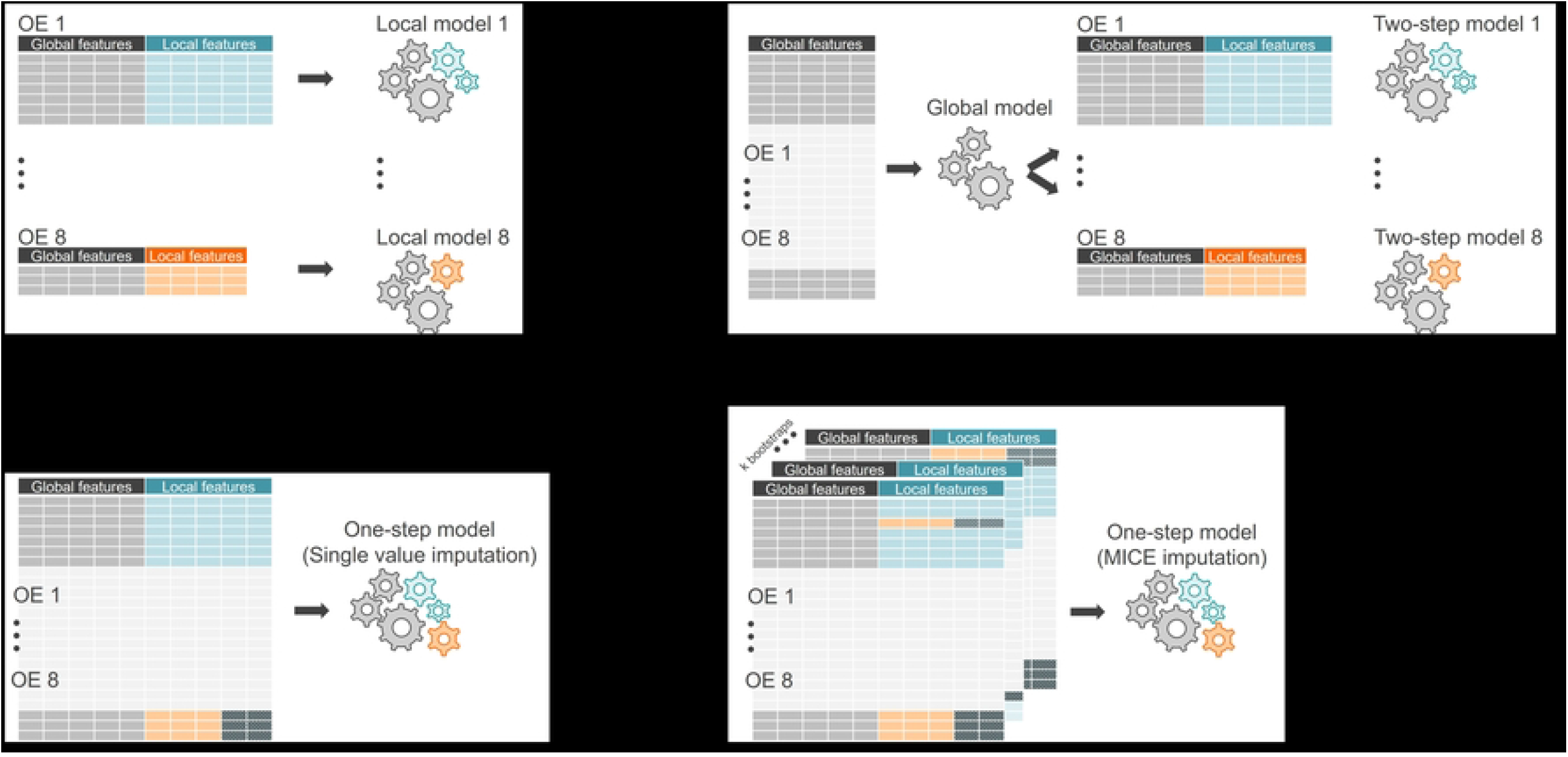
Comparison of the benchmarked models and their frameworks, with gearwheels representing the features.Grey stands for global features, blue and orange for local features specific to different countries, and patterned dark cells indicate missing values. A. Local model. B. Two-step model. C. One-step model with single value imputation. D. One-step model with MICE.

1. Local models for individual countries: For each country, we take this country’s data and run the model separately. This is, of course, only applicable if we have enough claims and exposure available for a given country as a solid foundation for training. The information contained in the each other countries about certain features and their correlation patterns to mortality rates remain unseen for each model.
2. Two-step approach: As detailed in the previous section, this approach combines global features in the first step model, using common features across countries. In the second step, a Local model is trained to capture also each country’s specificities based on residuals from the first step.
3. Global one-step with single value imputation: All datasets from different countries are combined in this early data fusion technique. The discrepancy in feature sets and values across countries results in missing blocks, as shown in Table 2.

**Table 2.** Percentage of missing values in each feature by country. For all three model types, missing values are imputed based on feature type: categorical features receive “Missing” and metric features receive “-1”. This approach retains information from non-missing values and identifies missing values during interactions for local features. In contrast, global features are free from missing values due to the design of the data collection process. In cases where a local model cannot be trained due to small data size, the One-Step approach may be the only viable option, but it results in missing blocks that must be imputed. The Two-Step model offers a valuable alternative by providing flexibility: if a local feature is entirely missing, it can be dropped, similar to local models, while global features are retained based on global patterns. For partially missing local features, single value imputation is applied, and the researcher has the option to drop or keep the imputed feature for a specific country. We chose to retain all features that are not completely missing within a country to ensure no information is lost.
4. Global one-step with bootstrapped multiple imputation: Similar to the previous approach, this method involves early data fusion by combining datasets from all countries. In this case, we use Bootstrapped Multiple Imputation with Decision Tree as imputation technique for missing values that arise due to the synthetic dataset creation. The procedure is as follows:
  - First draws k bootstrap samples from the combined dataset including missing values.
  - Fit a classification or regression tree by recursive partitioning, variable by variable.
  - After fitting a tree for the missing value based on the other values of the variable from the corresponding leaf, a value is randomly drawn.

| Country | F1 | F2 | F3 | F4 | F5 | F6 | F7 | F8 | F9 | F10 | F11 | F12 | F13 | F14 | F15 | F16 | F17 | F18 | F19 | F20 | F21 | F22 | F23 | F24 | F25 | F26 |
| --- | --- | --- | --- | --- | --- | --- | --- | --- | --- | --- | --- | --- | --- | --- | --- | --- | --- | --- | --- | --- | --- | --- | --- | --- | --- | --- |
| 1 | 0 | 0 | 0 | 0 | 0 | 0 | 0 | 0 | 0 | 0 | 0 | 33 | 0 | 72 | 0 | 0 | 0 | 0 | 0 | 0 | 0 | 72 | 72 | 72 | 72 | 0 |
| 2 | 0 | 0 | 0 | 0 | 0 | 0 | 0 | 0 | 0 | 0 | 0 | 4 | 0 | 0 | 0 | 0 | 0 | 0 | 0 | 0 | 48 | 48 | 48 | 48 | 48 | 48 |
| 3 | 0 | 0 | 0 | 0 | 0 | 0 | 0 | 0 | 0 | 0 | 0 | 0 | 0 | 0 | 0 | 33 | 33 | 0 | 0 | 0 | 0 | 33 | 5 | 6 | 0 | 0 |
| 4 | 0 | 0 | 0 | 0 | 0 | 0 | 0 | 0 | 0 | 0 | 0 | 28 | 0 | 0 | 0 | 28 | 0 | 28 | 0 | 0 | 0 | 28 | 0 | 28 | 28 | 28 |
| 5 | 0 | 0 | 0 | 0 | 0 | 0 | 0 | 0 | 0 | 0 | 0 | 4 | 0 | 0 | 0 | 0 | 0 | 0 | 0 | 0 | 0 | 72 | 0 | 100 | 100 | 0 |
| 6 | 0 | 0 | 0 | 0 | 0 | 0 | 0 | 0 | 0 | 0 | 0 | 37 | 0 | 0 | 0 | 62 | 0 | 0 | 0 | 0 | 62 | 58 | 56 | 0 | 62 | 0 |
| 7 | 0 | 0 | 0 | 0 | 0 | 0 | 0 | 0 | 0 | 0 | 0 | 0 | 0 | 0 | 8 | 2 | 8 | 0 | 6 | 0 | 8 | 8 | 8 | 8 | 8 | 8 |
| 8 | 0 | 0 | 0 | 0 | 0 | 0 | 0 | 0 | 0 | 0 | 0 | 0 | 0 | 6 | 0 | 6 | 0 | 0 | 0 | 6 | 6 | 0 | 0 | 6 | 6 | 6 |

This ensures that we can use it properly for multiple imputation, so that we are inducing some variation and not just the randomness in the leaf. The implementation was done in Python [30] with an adapted version of IterativeImputer [43], using 4 bootstrap samples and 2 imputations iterations each. We refer to [32] for further algorithm details. The number of iterations was determined based on a trial-and-error approach, as higher numbers had no significant impact on the final model results due to the dataset’s size. Based on each dataset resulting from the bootstrapped iteration, we trained the proposed model and finally pooled the eight predictions by averaging.

### Evaluation criteria

To evaluate our proposed methodology, we place a strong emphasis on two critical dimensions: predictive accuracy and computational efficiency. To gauge the predictive performance of our models, we employ two essential metrics: Root Mean Square Error (RMSE) for both in-sample and out-of-sample assessments. For a given country *j* it is calculated as follows:

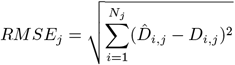

Additionally, we utilize the Poisson log-likelihood, which serves a dual role as a loss function and evaluation metric:

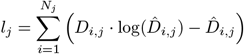

In the equations, 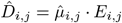 represents the predicted, while *D*_*i,j*_ the observed death counts. The in-sample metrics allow us to examine how well the model fits the training data. On the other hand, the out-of-sample metrics serves as a litmus test for the model’s ability to generalise to new, unseen data.

A higher log-likelihood and lower RMSE signify a closer fit between the model and the data, indicating superior performance. Conversely, a lower log-likelihood and higher RMSE are indicative of a less suitable model for the given data.

We consider runtime, memory usage, and storage requirements to evaluate the computational efficiency of our models, aiming for lower values to enhance their practical utility. These criteria offer a comprehensive assessment of our models’ performance in estimating mortality rates and pricing life insurance.

### Outcomes

This section details the benchmarking process for all four models, focusing on key metrics for performance and efficiency assessment. We evaluated the models using multiple metrics, including train and test RMSE and log-likelihood. Although RMSE is reported, log-likelihood is more reliable due to the distributional assumptions of the data. Additionally, we assessed computational efficiency through run time (seconds), memory consumption (megabytes), and storage space of the model object (kilobytes).

In Tables 3 and 4 we present the results exemplarily for country 5 and 7, and in an overview of all countries as well as the cross-country results. Each table provides an insight into the performance of the four benchmarked models, highlighting their strengths and weaknesses in various aspects. For ease of interpretation, we have used colour coding in dark grey to identify the best model within each row, based on the respective metric. The comparison is based on original values, before rounding for readability reasons.

**Table 3.** Performance evaluation for country 5.

| Metric | Local model | Two-step model | One-step model (Single Value) | One-step model (MICE) |
| --- | --- | --- | --- | --- |
| RMSE (Train) | $1.990 \times 10^{-2}$ | $1.979 \times 10^{-2}$ | $2.009 \times 10^{-2}$ | $2.126 \times 10^{-2}$ |
| RMSE (Test) | $1.709 \times 10^{-2}$ | $1.706 \times 10^{-2}$ | $1.709 \times 10^{-2}$ | $1.811 \times 10^{-2}$ |
| Log Likelihood (Train) | $-1.110 \times 10^4$ | $-1.069 \times 10^4$ | $-1.295 \times 10^4$ | $-1.315 \times 10^4$ |
| Log Likelihood (Test) | $-3.429 \times 10^3$ | $-3.399 \times 10^3$ | $-3.938 \times 10^3$ | $-3.998 \times 10^3$ |
| Runtime (Sec) | $1.370 \times 10^4$ | $3.970 \times 10^2$ | - | - |
| Memory (MB) | $2.998 \times 10^3$ | $1.663 \times 10^2$ | - | - |
| Storage (KB) | $2.174 \times 10^6$ | $2.162 \times 10^6$ | - | - |

**Table 4.** Performance evaluation for country 7.

| Metric | Local model | Two-step model | One-step model (Single Value) | One-step model (MICE) |
| --- | --- | --- | --- | --- |
| RMSE (Train) | $5.358 \times 10^{-2}$ | $5.061 \times 10^{-2}$ | $5.736 \times 10^{-2}$ | $5.847 \times 10^{-2}$ |
| RMSE (Test) | $3.542 \times 10^{-2}$ | $3.604 \times 10^{-2}$ | $3.983 \times 10^{-2}$ | $3.714 \times 10^{-2}$ |
| Log Likelihood (Train) | $-1.821 \times 10^3$ | $-1.469 \times 10^3$ | $-2.439 \times 10^3$ | $-2.682 \times 10^3$ |
| Log Likelihood (Test) | $-5.615 \times 10^2$ | $-5.529 \times 10^2$ | $-5.682 \times 10^2$ | $-5.693 \times 10^2$ |
| Runtime (Sec) | $9.144 \times 10^2$ | $1.518 \times 10^1$ | - | - |
| Memory (MB) | $3.041 \times 10^2$ | $1.983 \times 10^2$ | - | - |
| Storage (KB) | $9.816 \times 10^4$ | $9.376 \times 10^4$ | - | - |

Our Two-step modeling approach demonstrates the best predictive performance for nearly all countries, as evidenced by our comprehensive evaluation. This method outperforms Local models in most cases and shows significant advantages over the MICE method. Detailed results can be found in the tables and figures, highlighting the effectiveness of our approach.

The Two-step model shows the most substantial improvements for smaller countries (e.g., countries 7 and 8), compared to larger countries (e.g., countries 4 and 5). This is particularly evident in the test log-likelihood improvements from Local models to the Two-step model. By leveraging a Global model in the first step, we protect local specifics while enhancing the generalization capability, especially for smaller datasets.

Our research compares also one-step models, including single value imputation and MICE, with the proposed two-step approach. The findings consistently show that one-step models underperform and demand substantial computational resources. Specifically, MICE exhibits inferior performance for country-specific results. In terms of storage, single value imputation slightly outperforms the proposed model, if considered both steps. However, the one-step approaches require full retraining when new data becomes available, which can impact results for other countries.

When considering computational efficiency, encompassing aspects like runtime and memory consumption, the two-step approach stands out as the preferred choice. It’s important to emphasise that the performance of Local models is closely linked to the availability and quality of data within a given country. While this study has the privilege of using high-quality data with rich claims and exposures, this may not be the case for every country or data source. In such cases, the two-step approach with its cross-country learning capabilities provides a distinct advantage, as we can use the insights gained from the Global model to retrain the second step of the process.

Overall, our proposed two-step hierarchical modeling approach achieves superior predictive performance for nearly all countries, outperforming Local models and the MICE method, with log-likelihood proving to be a more reliable measure than RMSE due to the distributional assumptions of the data generation process. The Two-step model significantly enhances generalization for smaller countries, such as countries 7 and 8, by leveraging a Global model in the first step, which protects local specifics and improves performance even stronger compared to larger countries like countries 4 and 5.

## Summary and outlook

This study introduces a novel two-stage hierarchical mortality model that integrates global and local data to improve regional mortality risk estimation, particularly in data-scarce regions. The model leverages a LightGBM [31] in the first stage to capture global patterns, followed by country-specific refinements in the second stage. This approach demonstrated superior predictive accuracy compared to traditional methods and effectively addressed challenges related to missing data, scalability, and overgeneralization, offering a robust solution for mortality risk modeling across diverse regions.

The two-stage hierarchical modeling approach not only enhances predictive performance but also offers practical benefits in fields such as life insurance pricing, risk assessment, and public health planning. By generating more accurate mortality risk estimates, particularly in regions with limited local data, the model supports better-informed decision-making in industries that rely on precise risk evaluations. Its scalability and computational efficiency make it especially valuable in large-scale, multi-regional contexts.

Our model also stands out for its computational efficiency, excelling in runtime, memory usage, and storage requirements, particularly when the first-stage global model is excluded. This efficiency is advantageous when scaling to new countries, as only the second step requires retraining, leaving existing predictions unaffected. The reduced model size speeds up training times while maintaining high performance, making it suitable for applications where rapid training is essential. Additionally, the model provides an efficient solution for handling missing data, outperforming other methods like single-value imputation or MICE, particularly when working with small datasets where local data alone is insufficient, and the pre-learned knowledge of a larger model becomes critical.

Despite its strong performance across multiple regions, the model’s effectiveness depends on the availability and quality of data. In regions with low or inconsistent data quality, future research could explore more advanced imputation techniques or alternative methods for managing missing data. Further work could also investigate optimizing computational efficiency for even larger datasets or extending the model’s applicability to domains such as epidemiological forecasting, financial risk modeling, or public health surveillance. Integrating techniques like deep learning could enhance performance for more complex datasets, though this may compromise its interpretability.

The flexibility and robustness of the proposed hierarchical model open up new possibilities for accurate risk estimation, particularly in data-scarce environments. As industries continue to rely on precise mortality estimates for strategic decision-making, this approach sets the foundation for more reliable, scalable, and adaptable models capable of addressing the complexities of regional variability without compromising performance.

## Data Availability

The data are owned by a third party (insurance company) and authors do not have permission to share the data.

## Supporting information

**S1 Appendix. Rest of country-specific results**

**S2 Appendix. Hyperparameter optimization**

**S3 Appendix. Evaluation of prediction calibration**

## Notes

### Competing Interest Statement

The authors have declared no competing interest.

### Funding Statement

The author(s) received no specific funding for this work.

### Author Declarations

Data for the study was collected in a pseudonymised form from eight different operating units of a global primary insurance company, each representing a distinct country.

